# Parent and Adolescent Agreement on Ratings of ADHD Symptoms

**DOI:** 10.64898/2026.10.02.26364585

**Authors:** Steven W. Evans, Shreya Kupur Bhaskar, Emily Rosenthal, Marissa McClellan, Ross Boley, Eric A. Youngstrom

## Abstract

**Objective:** Parent-adolescent disagreement in ratings of attention-deficit/hyperactivity disorder (ADHD) symptoms can complicate clinical assessment. This study examined parent-adolescent agreement in ADHD symptom ratings and characteristics associated with disagreement.

**Method:** Participants were 1,008 parent-adolescent dyads recruited from a nationally stratified U.S. community sample. Adolescents were 12–18 years old (49.6% female) and along with their parents completed measures of ADHD and depressive symptoms. Agreement on Inattention (IN), Hyperactivity/Impulsivity (HI), and Total ADHD scores was examined. Item-level agreement was assessed using weighted kappa, Gwet’s AC2, percent agreement, and polychoric correlations. Associations between age, sex, and severity of ADHD and depression with levels of agreement were analyzed.

**Results:** Agreement for ADHD factor and total symptom scores was moderate (intraclass correlations = 0.67–0.71), with moderate-to-strong rank-order correspondence. Adolescents reported slightly higher ADHD symptom levels than parents, although the mean difference was small. Item-level agreement was moderate to high (Gwet’s AC2 = 0.79–0.95). Several HI items demonstrated the kappa’s paradox, with relatively low kappa values despite high Gwet’s AC2 and percent agreement. Q-correlations revealed low agreement on relative strengths and weaknesses. Age and sex were not associated with disagreement and parent-reported ADHD severity and adolescent-reported depressive symptoms were associated with disagreement.

**Conclusions:** Parent and adolescent ratings showed moderate agreement across boys and girls ages 12–18; however, disagreement related to relative strengths and weaknesses. Disagreement was associated with ADHD and depressive symptom severity. Findings suggest that ratings from parents and adolescents offer unique perspectives on the assessment of ADHD symptoms.

---

Attention Deficit-Hyperactivity Disorder (ADHD) is a neurodevelopmental disorder characterized by age-atypical levels of inattention and/or hyperactivity and impulsivity beginning early in life, causing substantial impairment, and presenting in multiple settings (APA, 2022). Declines in hyperactive/impulsive symptom severity over time are common; however, inattentive symptoms often persist (to a fluctuating extent) into adolescence and beyond (Sibley et al., 2022). During adolescence ADHD is associated with academic underperformance (Kent et al., 2011), social challenges (McKay et al., 2023; Smart et al., 2026), and emotion regulation difficulties (Astenvald et al., 2025), with higher risk for non-suicidal self-injury (Smanitto et al., 2026), risky driving (McManus et al., 2024; Narad et al., 2018), and substance use/abuse (Molina et al., 2013). It is important to highlight that those with subthreshold ADHD symptoms, even those who do not meet criteria for the disorder, are still at risk for negative outcomes relative to individuals with lower levels of such symptoms (e.g., Balázs & Keresztény, 2014; Hong et al., 2014).

ADHD symptoms are often assessed using standardized rating scales that correspond to DSM criteria for the disorder (such as the Vanderbilt, ADHD Rating Scale). These are typically completed by parents and teachers for children but regularly include self-reports for adolescents and adults (e.g., Eng et al., 2024). The gold standard diagnostic practice for ADHD is to obtain information from multiple reporters of ADHD symptoms and impairment, alongside clinical interviews (Bunford et al., 2026). Although there are concerns about cross-informant discordance between raters (e.g., Evans et al., 2005; Morris et al., 2021; Van Der Oord et al., 2006; Yeguez et al., 2016), ratings from multiple sources may capture important cross-situational differences in functioning and behavior (Eng et al., 2024; Sibley et al., 2012).

There are questions about the usefulness of self-reports of symptoms and impairment from adolescents, and this concern is partly based on a lack of agreement between adolescents and their parents. Adolescents with ADHD often under-report their symptoms and impairment relative to parent and teacher ratings (Fefer et al., 2018; Smith et al., 2000). For example, Sibley et al. (2019) found that two-thirds of a sample of adolescents with ADHD ages 11-15 years underreported symptoms relative to their parents, while nearly one-quarter denied all symptoms. Bivariate correlations between parent and adolescent factor scores for inattention (IA) and hyperactivity/impulsivity (HI) were statistically significant, but modest (0.23 & 0.33 respectively). Given the modest correlations, absolute agreement between parent and adolescent ratings was likely limited. Nevertheless, approximately one-third of parent-adolescent dyads agreed on the adolescent’s level of symptoms, highlighting substantial variability in cross-informant agreement. Smith and colleagues (2000) compared the symptom ratings of adolescents with ADHD to observation data and found very small relationships suggesting poor self-appraisal on the part of the adolescents. Colomer and colleagues (2020) found that although adolescents with ADHD reported fewer symptoms than their parents, their self-reported ratings of symptoms and impairment were still higher than those of peers without ADHD.

Inaccuracy in children and adolescents reporting of ADHD symptoms and impairment have been conceptualized as a positive illusory bias (PIB) attributed to youth with ADHD (Owens et al., 2007). Although many youths with ADHD may demonstrate a PIB in their reporting, as noted above this bias (or inaccuracy) is not uniform across all youth with ADHD. In fact, there is evidence that raises doubts about the PIB being associated with ADHD (Bourchtein et al., 2017; Watabe et al., 2018), suggesting that PIB may be characteristic of children with lower-than-average levels of competence. This raises the possibility that many youth perceive themselves as normal and view others in relation to themselves. Given the impairment of youth with ADHD, they are in a position of demonstrating a PIB when asked questions related to their symptoms and functioning.

Rather than assuming systematic bias in adolescent self-report, discrepancies may reflect differences in the information available to each reporter (De Los Reyes et al., 2013). As parent monitoring diminishes during the teenage years, adolescents have unique knowledge of a much greater portion of their day than a parent. Many adolescents with ADHD may exhibit fewer symptoms during the time outside of parent monitoring compared to when they are with their parents. This is certainly possible especially in relationships with substantial parent-child conflict. Although parent/adolescent disagreement may reflect biases in adolescent ratings, it may also reflect differences in the information available to each reporter. Consequently, clinicians are often left to rely on a variety of unclear factors when evaluating the credibility of informants and interpreting conflicting ratings (Youngstrom et al., 2011).

Evidence suggests a variety of reasons for this disagreement between parents and teens including the age and sex of the adolescent, and a teen’s level of depression. There is evidence that parent-adolescent disagreement for adolescents with ADHD in ratings of ADHD symptoms is larger for male than female adolescents (Frick et al., 2025) which is consistent with findings of greater self-awareness in adolescent girls than boys (Sharma et al., 2026). In a review of parent-child agreement on ratings of a variety of health dimensions, Hemmingsson et al., (2017) found mixed results pertaining to the effect of child age and gender with a similar number of studies reporting an effect with those reporting no effect. Part of mixed findings may be due to variability in sample sizes across the 39 studies (ranged from N=22 to N=742 with two exceptions (1,916 & 2,153) and 19 with samples less than 100) and the variety of constructs measured (e.g., pain, various psychiatric conditions, autonomy). Inconsistent findings in samples of youth with ADHD as well as samples without the disorder leave the effects of age and sex unclear.

In addition to the potential effect of age and sex, smaller discrepancies between parents’ and adolescents’ ratings were found for youth with ADHD who had high levels of depression symptoms compared to youth with ADHD without high levels of depression (Colomer et al., 2020; Jiang & Johnston, 2014, Swanson et al., 2012). In a review of these findings, Jiang and Johnston (2014) reported that the consensus of the literature was that depression symptoms among children diagnosed with ADHD may reduce these children’s overestimations of their competence. The study by Colomer and colleagues (2020) extended this finding to adolescents with ADHD. Both sets of authors noted that much of the research on this topic is limited due to investigators’ use of difference scores between parent’s and adolescent’s ratings as an index of agreement (Laird & De Los Reyes, 2013). Research is needed using alternative approaches that can accurately evaluate the relationship between depression and agreement.

Despite these concerns and mitigating factors; in regular clinical practice, it is often the adolescent and one parent who report youth symptoms, and the reliance on self-report increases during adolescence. First, as noted above, as children age an increasingly large portion of time is unsupervised by adults. Thus, adolescents are uniquely familiar with their own behavior in those situations. Second, many clinicians want ratings from a source in addition to a parent and teacher ratings are often less feasible and useful for adolescents than they are for children. Elementary school teachers know their students well and observe them in a variety of structured and unstructured activities; however, this is not true for adolescents. Middle and high school teachers may have only superficial relationships with many of their 100 or more students. In addition, they typically see any individual student for less than an hour each day and only in a very specific context. Third, there can be substantial variability in how a student performs and behaves across classes due to variability in the student’s ability in the subject matter and how much they like the teacher. Thus, it is not surprising that agreement on symptom and functioning rating scales between secondary school teachers is poor for adolescents with ADHD (Evans et al., 2005) and it is unknown to what extent variability is due to actual differences in student behavior between classes or error. Finally, from a clinician’s perspective, there is also difficulty obtaining completed rating scales from middle and high school teachers as return rates can be quite poor (e.g., DuPaul et al., 2021). As a result, self-ratings are the most convenient and practical source for ratings in addition to parents.

It is important to note that the majority of the findings described above are restricted to disagreement between ratings by youth with ADHD and others (Volz-Sidiropoulou et al., 2016). Findings about agreement specific to youth with ADHD may or may not apply to adolescents presenting for an evaluation or being screened for the disorder as some portion of both groups do not have ADHD. Understanding agreement on ratings of ADHD symptoms across all adolescents and considering the potential role of the severity of ADHD symptoms can provide a clearer picture of agreement.

Our limited understanding of parent-adolescent agreement when rating symptoms of ADHD may be partly due to how prior studies defined and investigated interrater (dis)agreement in reports of ADHD symptoms. Inter-rater agreement between parents and adolescents has often been assessed using discrepancy or difference scores (e.g., subtracting the score of one rater from that of another), Cohen’s Kappa, and/or Intraclass Correlation Coefficients (ICCs). Researchers have highlighted that using difference scores provides poor model fit for observed data (De Los Reyes et al., 2013; Sibley et al., 2019) and may not serve as a valid indicator of informant discrepancies (De Los Reyes et al., 2013). The use of discrepancy scores increases the likelihood of Type I errors due to, for example, unequal variability across raters and/or bivariate associations between variables (De Los Reyes et al., 2013; Sibley et al., 2019). Despite some strengths, concerns have also been raised about the possibility for biased Weighted Cohen’s Kappa estimates of interrater reliability in the context of skewed ordinal data. Specifically, Cohen’s Kappa estimates can be biased by distributional skew and the dissimilarity in category frequency across raters (Wongpakaran et al., 2013). These features can lead to paradoxically low kappa value despite high percentage agreement between raters (“kappa paradox”, Wongpakaran et al., 2013).

One alternative to avoiding the kappa paradox when calculating interrater reliability for ordinal data is Gwet’s AC2 (Gwet, 2014). This statistic reflects the conditional probability of agreement between two raters after correcting for the possibility of raters agreeing due to chance (Wongpakaran et al., 2013). In the context of ordinal data with low prevalence, this statistic has shown high accuracy in predicting and approximating interrater reliability in simulations (**Zhao et al., 2022**) and is less prone to bias due to skewed distribution or imbalances across categories and across raters than traditional kappa statistics (Wongpakaran et al., 2013). Therefore, Gwet’s AC2 may be a better indicator of parent-child agreement than kappa on individual items given the known limitations of the kappa statistic.

In summary, previous research raised questions about the validity of adolescent self-reports of ADHD symptoms due to disagreement between parent and child ratings. Child characteristics associated with disagreement suggest that sex, age, and level of depression may impact agreement. In addition, there is evidence that severity of ADHD symptoms may impact agreement as levels of agreement between parents and adolescents are worse for youth with ADHD than those without. As a clinician does not initially know if an adolescent presenting for an evaluation meets criteria for ADHD or not, it is important to examine agreement across youth with a wide range of symptoms. Further, clinicians often have limited options for obtaining symptom ratings from people other than the parent and adolescent so understanding their likelihood of agreement and characteristics that may influence agreement are important. Understanding agreement for factor scores and individual items across a large diverse sample can advance our ability to interpret assessment data. For instance, if an item displays uniquely high levels of disagreement compared to other items, it may warrant interpretation relative to base rates, and clinicians should think critically about potential biases on specific items when interpreting self-report data. In addition, advanced analytical approaches other than difference scores and Cohen’s Kappa are needed to improve our ability to quantify degrees of agreement on ADHD rating scales.

## Current Study

In this study we examined parent-adolescent agreement on ratings of ADHD symptoms for adolescents 12 to 18 years of age and their parents. This is a large (N=1,008) and diverse community sample and we examined levels of agreement on factor scores and individual items. We examined degrees of agreement in relation to demographic characteristics as well as clinical characteristics. Analyses were conducted using multiple advanced techniques including Gwet’s AC2 for the examination of agreement with individual items and kappa to examine agreement with factor scores. Results were compared across agreement indices to screen for evidence of “kappa’s paradox” and examine the extent to which Gwet’s AC2 is resistant to that effect. Specifically, we anticipated that given the skewed data resulting from this measure, items with a high percent agreement may have low kappas, but Gwet’s AC2 indices will be commensurate with the percent agreement data.

The research questions are, 1) To what degree is there agreement in ratings of ADHD symptoms between parents and adolescents on ADHD factor scores and individual items? 2) To what extent are child age, sex, and levels of ADHD and depression symptoms associated with levels of agreement on the scales?

## Method

### Participants

The sample consisted of 1,008 parents of a 12-to 18-year-old adolescents (*M_age_* = 47.4 years, *SD* = 8.8; 72.0% female) and 1,008 of their adolescent children aged 12–18 years (*M_age_* = 14.4 years, *SD* = 1.6; 49.6% female). The parent-child dyads were recruited from all 50 states of the United States of America and the District of Columbia (see Table 1).

**Table 1.** Parent and Adolescent Characteristics.

|  |  |
| --- | --- |
| ***Youth Characteristics (n = 1,008)*** |  |
| Child age, *M (SD)* | 14.4 (1.6) |
| Age, *n (%)* |  |
| 12 | 163 (16.2%) |
| 13 | 179 (17.8%) |
| 14 | 165 (16.4%) |
| 15 | 190 (18.8%) |
| 16 | 192 (19.0%) |
| 17 | 109 (10.8%) |
| 18 | 10 (1.0%) |
| Sex, *n (%)* |  |
| Male | 506 (50.5%) |
| Female | 500 (49.6%) |
| Intersex/Other | 2 (0.2%) |
| ***Parent Characteristics (n = 1,008)*** |  |
| Parent age, *M (SD)* | 47.4 (8.8 years) |
| Range (Median) | 25 – 81 (46 years) |
| Gender, *n (%)* |  |
| Male | 274 (27.2%) |
| Female | 726 (72.0%) |
| Non-binary or Other | 8 (0.8%) |
| Race, *n (%)* |  |
| White | 842 (83.5%) |
| Black or African American | 93 (9.2%) |
| Asian | 29 (2.9%) |
| Native American | 32 (3.2%) |
| Native Hawaiian or Pacific Islander | 3 (0.3%) |
| Other | 38 (3.8%) |
| Prefer not to say | 12 (1.2%) |
| Ethnicity – Hispanic or Latino, *n (%)* | 113 (11.2%) |
| Education, *n (%)* |  |
| High school or less | 158 (15.7%) |
| Some college | 231 (22.9%) |
| 2-year, 4-year, or post graduate degree | 609 (61.4%) |
| Marital Status – Married, *n (%)* | 642 (63.7 %) |
| Family Income, *n (%)* |  |
| < \$10,000 | 39 (3.9%) |
| \$10,000 – 39,999 | 174 (17.3%) |
| \$40,000 – 69,999 | 213 (21.0%) |
| \$70,000 – 99,999 | 191 (18.9%) |
| > \$100,000 | 352 (34.9%) |
| U.S. Region, *n (%)* |  |
| Northeast | 195 (19.3%) |
| Midwest | 236 (23.4%) |
| South | 386 (38.3%) |
| West | 191 (18.9%) |

### Procedure

Participants were recruited through YouGov, an international online survey company, for the Nationwide Children’s Hospital’s Behavioral Evaluation and Norms for Child Health and MARKers of Well-being (BENCHMARK) Project—a team of investigators collecting normative data for several parent-and youth-report measures assessing mental and behavioral health, academic performance, and other related constructs. YouGov collected and cleaned all survey responses, paired parent and adolescent data to create dyads, and applied stratification procedures based on key demographics (child sex, race, ethnicity, and parent education level) resulting in a nationally stratified U.S. sample. All study procedures were reviewed and designated Not Human Subjects Research by the Institutional Review Board at Nationwide Children’s Hospital. Data collection occurred between July 11, 2025, and March 7, 2026.

### Measures

#### Vanderbilt Assessment Scales (VAS; Wolraich et al., 2003)

The VAS is a 55-item parent-report scale used to identify symptoms of ADHD and other co-morbid disorders, and levels of performance in youth over a 6-month period. The VAS was written as a parent-report scale, so instructions and items were reworded to be grammatically appropriate for a youth respondent without changing the item content. Parents and youth completed 18 items from this scale covering all 18 *DSM-IV* ADHD symptoms (9 inattentive, 9 hyperactive/impulsive) with higher ratings reflecting more severity. Items are scored on a 4-point Likert scale (0: Never, 1: Occasionally, 2: Often, 3: Very Often). The parent version of the scale used in this study is freely available at the National Institute for Children’s Health Quality (NICHQ) website (https://nichq.org/wp-content/uploads/2024/09/NICHQ-Vanderbilt-Assessment-Scales.pdf).

#### Very Quick Inventory of Depressive Symptomatology (VQIDS; self and parent report)

Parent-and youth-reported depressive symptoms were assessed using a 5-item depression symptom scale. The items ask about sadness, concentration and decision-making difficulties, negative self-evaluation, loss of interest, and low energy levels during the past 7 days. Items are scored on a 4-point ordinal scale ranging from 0-3, with higher scores indicating greater depressive symptom severity. The VQIDS has demonstrated good to excellent internal consistency and validity in relation to more extensive measures of depression (Croarkin et al., 2025).

### Statistical Analysis

To address the first research question, ICC using a two-way random-effects model with absolute agreement and single measures, ICC (2,1), were calculated to assess agreement between parent-and youth-reported IA and HI factor scores and Total ADHD scores (Shrout & Fleiss, 1979; Koo & Li, 2016). ICCs assess absolute agreement in the actual magnitude of scores, indicating the extent to which parent and youth ratings are similar in level. We used Koo and Li (2016) guidelines for classifying ICC indices (< 0.5=poor; 0.5-0.75=moderate; 0.75-0.9=good; >0.9=excellent). Spearman correlations were included to provide complementary information about rank-order correspondence for factor scores and the total score. Q-correlations were used to assess the extent to which parents and children identify the same relative pattern of symptoms across items, providing an index of profile correspondence that is distinct from agreement in overall symptom severity or rank ordering. Mean Q-correlations were calculated for IA, HI, and Total ADHD symptoms.

Parent-youth differences in IA and HI factor scores and Total ADHD scores were evaluated using paired comparisons, and the standardized magnitude of the within-participant difference was quantified using Cohen’s dz (Lakens, 2013).

For each of the 18 Vanderbilt ADHD symptom items, parent-youth agreement was evaluated using multiple methods. Weighted Cohen’s κ was used to assess agreement while accounting for the ordinal nature of the item-response categories (Cohen, 1968). Because Cohen’s κ can be affected by skewed or imbalanced response distributions, Gwet’s AC2 was calculated as an alternative agreement coefficient (Gwet, 2008, 2014). Percent exact agreement was calculated to describe the proportion of parent-youth item ratings that were identical. Finally, Polychoric correlations were calculated to evaluate the association between parent and youth item ratings while accounting for the ordinal response structure (Olsson, 1979).

To examine the first part of the second research question regarding the impact of age and sex on agreement, analyses were stratified. Age was categorized into younger adolescents (12-14 years) and older adolescents (15-18 years), and analyses were conducted separately for males and females. Item-level agreement was examined within each age-by-sex subgroup using weighted Cohen’s κ, Gwet’s AC2, and percent exact agreement. Descriptive statistics, including means, standard deviations, and skewness, were calculated for individual item scores to characterize the distribution of responses within these subgroups. The second part of this research question pertained to the relationship between severity of symptoms of ADHD and depression with indices of parent-adolescent agreement. Bland-Altman analyses (modified Bland-Altman for depression scores) were conducted to examine this relationship.

Statistical significance was evaluated using a two-sided αlevel of 0.05. All analyses were conducted in R (v4.5.2) using RStudio (v2026.01.1).

## Results

### Research Question 1: To what degree is there agreement in ratings of ADHD symptoms between parents and adolescents on ADHD factor scores and individual items?

At the factor and total level, parent-youth agreement was moderate. The ICC (2,1) was 0.67 (95% CI [0.64,0.71]) for the IA factor score, 0.69 (95% CI [0.65,0.72]) for the HI factor score, and 0.71 (95% CI [0.68,0.74]) for Total ADHD symptoms. These ICC scores are considerate moderate agreement (Koo & Li, 2016). Spearman correlations were similar, ranging from ρ= 0.67 to 0.71, indicating moderate-to-strong rank-order correspondence between parent-youth ratings. Youth reported slightly higher total scores than parents, with mean scores of 11.67 and 11.13, respectively, corresponding to a mean parent-youth difference of −0.54 points. Although this difference was statistically significant, its standardized magnitude was small (Cohen’s dz = −0.07; see Table 2). Thus, at the group level there is evidence of overall agreement on the total score and factors scores.

**Table 2.**
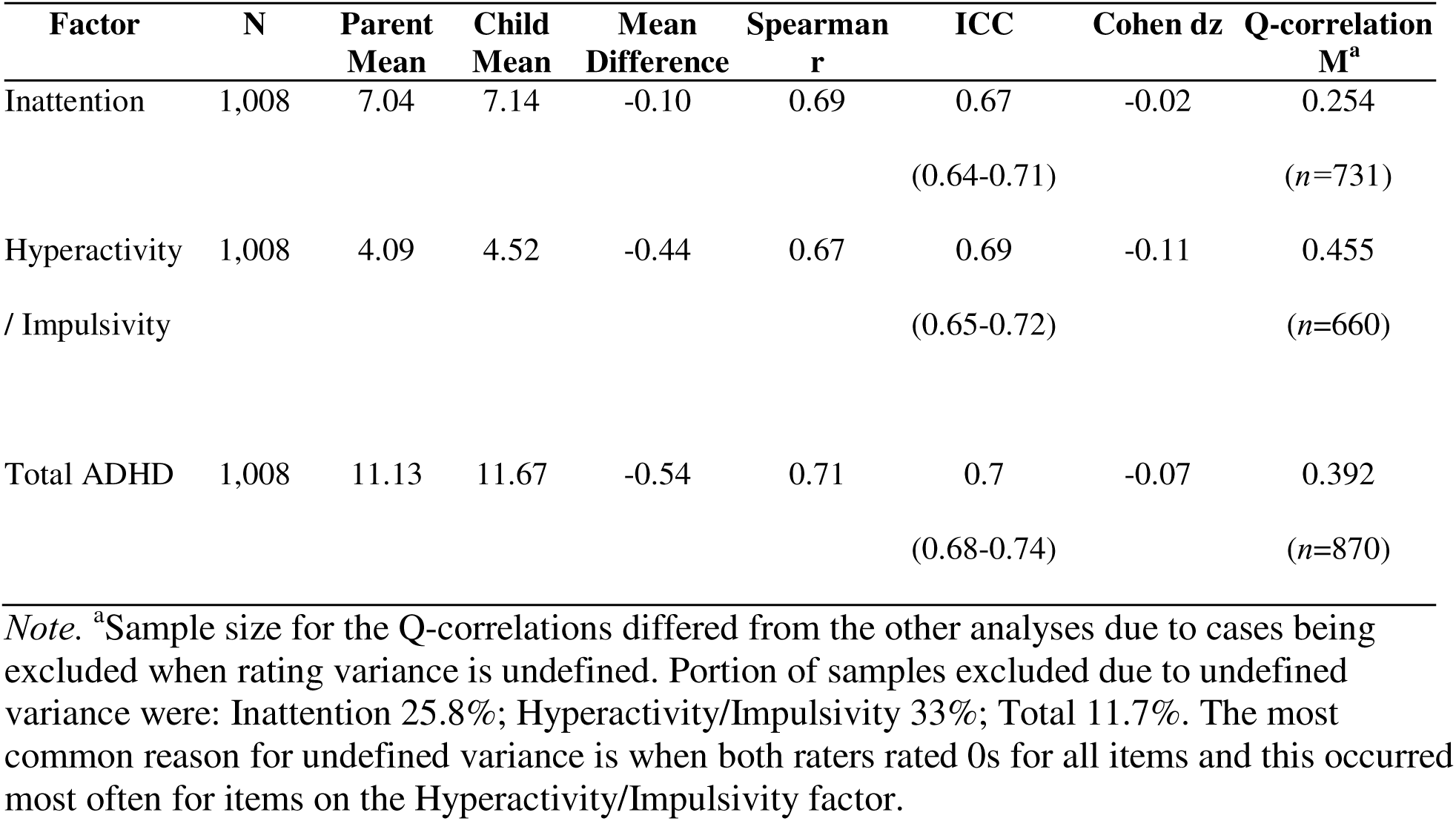
Parent-Adolescent Agreement Across ADHD Factors.

| **Factor** | **N** | **Parent**<br>**Mean** | **Child**<br>**Mean** | **Mean**<br>**Difference** | **Spearman**<br>**r** | **ICC** | **Cohen dz** | **Q-correlation**<br>**M<sup>a</sup>** |
| --- | --- | --- | --- | --- | --- | --- | --- | --- |
| Inattention | 1,008 | 7.04 | 7.14 | -0.10 | 0.69 | 0.67 | -0.02 | 0.254 |
|  |  |  |  |  |  | (0.64-0.71) |  | (n=731) |
| Hyperactivity | 1,008 | 4.09 | 4.52 | -0.44 | 0.67 | 0.69 | -0.11 | 0.455 |
| / Impulsivity |  |  |  |  |  | (0.65-0.72) |  | (n=660) |
| Total ADHD | 1,008 | 11.13 | 11.67 | -0.54 | 0.71 | 0.7 | -0.07 | 0.392 |
|  |  |  |  |  |  | (0.68-0.74) |  | (n=870) |

*Note.* <sup>a</sup>Sample size for the Q-correlations differed from the other analyses due to cases being excluded when rating variance is undefined. Portion of samples excluded due to undefined variance were: Inattention 25.8%; Hyperactivity/Impulsivity 33%; Total 11.7%. The most common reason for undefined variance is when both raters rated 0s for all items and this occurred most often for items on the Hyperactivity/Impulsivity factor.

The Q-correlations present a very different and complementary perspective on parent-child agreement. Mean Q-correlations were 0.254 for Inattention, 0.455 for Hyperactivity/Impulsivity, and 0.392 for Total ADHD. These values indicate that within individual children, parent and child reports showed modest correspondence in the relative pattern of symptoms endorsed. This was particularly true for Inattention. Despite a Spearman correlation of 0.69, an ICC of 0.67, virtually identical parent and child means, and a negligible mean difference (*d*z=-0.02), the mean Q-correlation was only 0.254. Thus, parents and children tended to agree about which children had relatively higher versus lower levels of inattentive symptoms and about the overall severity of inattention. However, they showed substantially less agreement regarding the specific pattern of inattentive symptoms characterizing an individual child.

Hyperactivity/Impulsivity showed the greatest within-person profile correspondence (mean Q = 0.455), although this was moderate. Total ADHD was intermediate (mean Q = 0.392). These findings indicate that traditional indices such as correlations and ICCs may suggest moderate to strong agreement, whereas Q-correlations reveal considerably less concordance in the symptom profiles reported for individual children. Consequently, the Q-correlations qualify and refine conclusions drawn from conventional measures of agreement by highlighting discrepancies in the specific symptoms endorsed, even when overall severity ratings are similar.

At the item level, parent-youth agreement across the 18 Vanderbilt item scores was generally moderate based on weighted Cohen’s κ, with values ranging from κ=0.42 to 0.63 (see Tables 3, 4, & 5). Gwet’s AC2 values ranged from 0.79 to 0.95, indicating high levels of agreement. Percent exact agreement ranged from 53.2% to 82%. Polychoric correlations ranged from 0.59 to 0.72, indicating moderate-to-strong associations between parent and youth item ratings.

**Table 3.** Item-level Parent-Child Agreement.

| VAS Items | N | Weighted<br>Kappa | Gwet AC2 | Percent<br>Agreement | Polychoric_r |
| --- | --- | --- | --- | --- | --- |
| 1: attention to details | 1005 | 0.52 | 0.85 | 57.41 | 0.62 |
| 2: sustaining attention | 1005 | 0.52 | 0.81 | 55.72 | 0.62 |
| 3: does not listen | 1006 | 0.48 | 0.83 | 58.05 | 0.59 |
| 4: follow through | 1007 | 0.49 | 0.83 | 59.98 | 0.60 |
| 5: difficulty organizing | 1007 | 0.50 | 0.79 | 55.51 | 0.59 |
| 6: avoids tasks | 1006 | 0.55 | 0.79 | 54.47 | 0.64 |
| 7: loses things | 1008 | 0.48 | 0.83 | 58.73 | 0.59 |
| 8: easily distracted | 1007 | 0.57 | 0.79 | 53.23 | 0.65 |
| 9: forgetful | 1005 | 0.53 | 0.82 | 58.21 | 0.63 |
| 10: fidgets | 1006 | 0.63 | 0.81 | 59.24 | 0.72 |
| 11: leaves seat | 1007 | 0.52 | 0.93 | 75.07 | 0.68 |
| 12: runs & climbs | 1008 | 0.42 | 0.95 | 82.04 | 0.64 |
| 13: playing quietly | 1008 | 0.46 | 0.93 | 73.21 | 0.64 |
| 14: on the go | 1003 | 0.50 | 0.90 | 69.09 | 0.65 |
| 15: talks too much | 1002 | 0.53 | 0.83 | 60.38 | 0.63 |
| 16: blurts answers | 1006 | 0.51 | 0.87 | 64.91 | 0.64 |
| 17: difficulty waiting | 1007 | 0.52 | 0.89 | 68.32 | 0.66 |
| 18: interrupts | 1008 | 0.53 | 0.87 | 65.77 | 0.66 |

*Note.* A few participants skipped items resulting in the variability in samples size indicated in the column labelled “N”.

**Table 4:** Inattention Item-level Parent-Youth Agreement by Age-sex sub-group.

| Age Group X<br>Sex | Item | N | Weighted<br>Kappa | Gwet's_AC2 | Percent<br>Agreement | Mean | SD | Skewness |
| --- | --- | --- | --- | --- | --- | --- | --- | --- |
| 12-14_Male | 8 | 286 | 0.63 | 0.83 | 56.64 | 1.02 | 0.90 | 0.73 |
| 15-18_Female | 8 | 265 | 0.59 | 0.79 | 54.34 | 0.85 | 0.92 | 0.94 |
| 12-14_Female | 6 | 234 | 0.59 | 0.79 | 58.97 | 0.76 | 0.92 | 1.03 |
| 12-14_Female | 9 | 235 | 0.58 | 0.83 | 58.30 | 0.75 | 0.84 | 1.03 |
| 15-18_Female | 5 | 266 | 0.58 | 0.85 | 63.91 | 0.67 | 0.80 | 1.15 |
| 12-14_Female | 2 | 235 | 0.56 | 0.81 | 57.02 | 0.80 | 0.86 | 0.98 |
| 15-18_Female | 6 | 266 | 0.56 | 0.81 | 56.77 | 0.69 | 0.88 | 1.22 |
| 12-14_Male | 6 | 286 | 0.56 | 0.78 | 52.80 | 0.98 | 0.93 | 0.71 |
| 12-14_Male | 3 | 286 | 0.55 | 0.83 | 55.59 | 0.77 | 0.88 | 1.12 |
| 15-18_Female | 7 | 266 | 0.55 | 0.86 | 63.53 | 0.57 | 0.77 | 1.47 |
| 12-14_Female | 8 | 235 | 0.54 | 0.77 | 49.79 | 0.96 | 0.87 | 0.68 |
| 12-14_Male | 1 | 286 | 0.53 | 0.84 | 54.55 | 1.04 | 0.80 | 0.72 |
| 15-18_Male | 9 | 220 | 0.53 | 0.84 | 56.36 | 0.71 | 0.83 | 1.12 |
| 12-14_Male | 9 | 285 | 0.53 | 0.80 | 57.54 | 0.88 | 0.85 | 0.81 |
| 12-14_Male | 2 | 284 | 0.52 | 0.80 | 53.17 | 0.98 | 0.87 | 0.78 |
| 15-18_Male | 4 | 219 | 0.51 | 0.85 | 64.38 | 0.67 | 0.83 | 1.23 |
| 12-14_Female | 1 | 235 | 0.51 | 0.85 | 54.04 | 0.82 | 0.75 | 0.74 |
| 15-18_Female | 9 | 264 | 0.51 | 0.82 | 60.61 | 0.71 | 0.83 | 1.22 |
| 15-18_Female | 4 | 266 | 0.51 | 0.87 | 64.66 | 0.52 | 0.73 | 1.56 |
| 15-18_Female | 1 | 264 | 0.50 | 0.88 | 63.64 | 0.75 | 0.74 | 1.06 |
| 15-18_Female | 2 | 266 | 0.50 | 0.81 | 55.64 | 0.72 | 0.85 | 1.17 |
| 12-14_Female | 5 | 235 | 0.50 | 0.77 | 53.62 | 0.72 | 0.84 | 1.06 |
| 15-18_Male | 1 | 219 | 0.49 | 0.84 | 57.53 | 0.90 | 0.83 | 0.96 |
| 15-18_Male | 2 | 219 | 0.49 | 0.81 | 57.53 | 0.87 | 0.85 | 1.01 |
| 15-18_Male | 8 | 220 | 0.49 | 0.79 | 51.36 | 0.86 | 0.85 | 0.86 |
| 12-14_Male | 4 | 286 | 0.48 | 0.80 | 55.94 | 0.81 | 0.84 | 0.83 |
| 15-18_Male | 3 | 219 | 0.48 | 0.83 | 57.53 | 0.69 | 0.84 | 1.19 |
| 15-18_Male | 6 | 219 | 0.48 | 0.77 | 49.32 | 0.86 | 0.90 | 0.96 |
| 12-14_Male | 7 | 286 | 0.48 | 0.82 | 57.34 | 0.83 | 0.80 | 0.98 |
| 15-18_Male | 5 | 219 | 0.48 | 0.79 | 55.71 | 0.84 | 0.90 | 1.05 |
| 12-14_Female | 7 | 235 | 0.48 | 0.82 | 54.47 | 0.69 | 0.76 | 0.99 |
| 15-18_Female | 3 | 266 | 0.47 | 0.87 | 63.16 | 0.51 | 0.73 | 1.47 |
| 12-14_Male | 5 | 286 | 0.46 | 0.74 | 48.95 | 0.10 | 0.90 | 0.65 |
| 12-14_Female | 4 | 235 | 0.45 | 0.80 | 55.75 | 0.58 | 0.79 | 1.27 |
| 15-18_Male | 7 | 220 | 0.39 | 0.83 | 59.09 | 0.66 | 0.75 | 1.24 |
| 12-14_Female | 3 | 234 | 0.39 | 0.80 | 55.56 | 0.60 | 0.79 | 1.26 |

**Table 5:** Hyperactivity/Impulsivity Item-level Parent-Youth Agreement by Age-sex sub-group.

| Age Group X Sex | Item | N | Weighted Kappa | Gwet's AC2 | Percent Agreement | Mean | SD | Skewness |
| --- | --- | --- | --- | --- | --- | --- | --- | --- |
| 12-14_Male | 10 | 285 | 0.65 | 0.77 | 57.90 | 0.94 | 1.01 | 0.78 |
| 15-18_Female | 10 | 266 | 0.62 | 0.83 | 60.15 | 0.64 | 0.85 | 1.30 |
| 12-14_Female | 10 | 235 | 0.61 | 0.78 | 58.72 | 0.76 | 0.90 | 1.12 |
| 12-14_Male | 11 | 286 | 0.60 | 0.91 | 72.73 | 0.43 | 0.74 | 1.88 |
| 15-18_Male | 10 | 219 | 0.60 | 0.85 | 60.73 | 0.67 | 0.85 | 1.20 |
| 12-14_Male | 17 | 286 | 0.58 | 0.87 | 65.04 | 0.55 | 0.77 | 1.41 |
| 15-18_Male | 15 | 219 | 0.58 | 0.89 | 64.84 | 0.56 | 0.74 | 1.34 |
| 12-14_Male | 12 | 286 | 0.57 | 0.94 | 79.72 | 0.30 | 0.61 | 2.27 |
| 12-14_Male | 14 | 285 | 0.57 | 0.88 | 62.81 | 0.55 | 0.80 | 1.49 |
| 12-14_Female | 18 | 235 | 0.57 | 0.88 | 73.62 | 0.53 | 0.75 | 1.46 |
| 15-18_Female | 16 | 266 | 0.55 | 0.89 | 66.17 | 0.44 | 0.70 | 1.77 |
| 15-18_Female | 15 | 266 | 0.55 | 0.82 | 62.03 | 0.65 | 0.79 | 1.07 |
| 12-14_Female | 17 | 234 | 0.54 | 0.90 | 67.52 | 0.37 | 0.69 | 2.14 |
| 15-18_Male | 18 | 220 | 0.52 | 0.89 | 64.55 | 0.49 | 0.73 | 1.58 |
| 15-18_Female | 18 | 266 | 0.51 | 0.88 | 66.17 | 0.42 | 0.71 | 1.95 |
| 12-14_Female | 14 | 233 | 0.51 | 0.92 | 73.82 | 0.31 | 0.64 | 2.37 |
| 12-14_Male | 15 | 284 | 0.51 | 0.82 | 57.04 | 0.74 | 0.82 | 1.02 |
| 12-14_Male | 18 | 286 | 0.51 | 0.82 | 59.79 | 0.73 | 0.85 | 1.07 |
| 12-14_Male | 16 | 284 | 0.50 | 0.83 | 58.10 | 0.59 | 0.81 | 1.38 |
| 12-14_Male | 13 | 286 | 0.50 | 0.91 | 67.83 | 0.42 | 0.70 | 1.68 |
| 12-14_Female | 16 | 235 | 0.50 | 0.89 | 67.66 | 0.39 | 0.66 | 1.82 |
| 15-18_Female | 11 | 265 | 0.48 | 0.95 | 80.38 | 0.23 | 0.58 | 2.83 |
| 15-18_Female | 17 | 266 | 0.47 | 0.89 | 72.56 | 0.36 | 0.71 | 2.16 |
| 12-14_Female | 11 | 235 | 0.47 | 0.93 | 76.60 | 0.23 | 0.56 | 2.69 |
| 15-18_Male | 14 | 219 | 0.50 | 0.89 | 70.32 | 0.42 | 0.78 | 2.04 |
| 12-14_Female | 15 | 232 | 0.50 | 0.79 | 58.19 | 0.70 | 0.79 | 1.08 |
| 15-18_Male | 17 | 220 | 0.45 | 0.89 | 68.18 | 0.47 | 0.72 | 1.79 |
| 12-14_Female | 13 | 235 | 0.45 | 0.92 | 72.34 | 0.29 | 0.66 | 2.75 |
| 15-18_Male | 13 | 220 | 0.44 | 0.93 | 75.00 | 0.30 | 0.57 | 2.09 |
| 15-18_Male | 11 | 220 | 0.43 | 0.90 | 70.00 | 0.38 | 0.72 | 2.01 |
| 15-18_Male | 16 | 220 | 0.43 | 0.89 | 69.09 | 0.40 | 0.65 | 1.58 |
| 15-18_Female | 13 | 266 | 0.42 | 0.95 | 78.20 | 0.21 | 0.54 | 2.94 |
| 15-18_Female | 14 | 265 | 0.41 | 0.90 | 70.94 | 0.32 | 0.63 | 2.08 |
| 12-14_Female | 12 | 235 | 0.33 | 0.96 | 82.55 | 0.12 | 0.41 | 4.24 |
| 15-18_Female | 12 | 266 | 0.31 | 0.97 | 84.96 | 0.13 | 0.41 | 3.60 |
| 15-18_Male | 12 | 220 | 0.29 | 0.94 | 81.36 | 0.20 | 0.54 | 3.38 |

Evidence of kappa’s paradox was notable in the results of the HI items, particularly Items 11-14 (see Table 5). These items generally demonstrated comparatively lower weighted κ values despite high Gwet’s AC2 and high percent exact agreement. For example, Item 12 had weighted κ values ranging from approximately 0.29 to 0.33 in some age-by-sex subgroups, and Gwet’s AC2 ranged from approximately 0.94 to 0.97 and exact agreement exceeded 79%. The discrepancy between weighted κ and Gwet’s AC2 should be interpreted in the context of the highly concentrated and positively skewed response distributions observed for these item scores. The response distributions for Items 11-14 were characterized by relatively low mean ratings and positive skewness. Item 12, for example, had mean ratings ranging from 0.12 to 0.30 and skewness values ranging from approximately 2.27 to 4.24 across age-by-sex groups. These findings indicate that responses to these items were concentrated at the lower end of the rating scale consistent with expected low levels of HI symptoms in adolescents. Despite relatively low κ estimates for some items, observed exact agreement and Gwet’s AC2 indicated high parent-youth correspondence. Thus, the lower κ estimates may reflect the influence of the marginal response distributions on chance-corrected agreement rather than substantively poor agreement between informants (Gwet, 2008, 2014).

Overall, agreement was stronger for summed ADHD factor and total scores than for individual item ratings or patterns of symptom endorsement, with the highest correspondence observed for the total score.

### Research Question 2: To what extent are the age and sex associated with levels of agreement on the scales and to what degree does agreement vary as a function of severity of depression and ADHD symptoms?

Parent-youth item level agreement was broadly similar across age and sex groups, although modest differences were observed (see Tables 4 & 5). Younger adolescents aged 12-14 years demonstrated a somewhat higher agreement for the total score than older adolescents aged 15-18 years. Among younger adolescents, ICC (2,1) was 0.73 and Spearman’s ρ was 0.74, compared with an ICC of 0.67 and ρ of 0.68 among older adolescents. Mean item-level weighted κ values were also somewhat higher among younger adolescents (approximately 0.53 vs. 0.49). However, these differences were modest.

Agreement was similarly consistent across sex. Males demonstrated an ICC of 0.70 and Spearman’s ρ of 0.69, whereas females demonstrated an ICC of 0.72 and ρ of 0.74. Mean item-level weighted κ values were nearly identical between males (κ=0.51) and females (κ=0.52). In summary, we did not find evidence of meaningful sex-related differences in overall parent-youth agreement.

Age-by-sex subgroup analyses showed the same general pattern. Total-score agreement was highest among females ages 12-14 years (ICC=0.74; ρ=0.75), followed by males aged 12-14 years (ICC= 0.73; ρ= 0.73), females aged 15-18 years (ICC=0.70; ρ=0.72), and males aged 15-18 years (ICC=0.64; ρ=0.63). All subgroup correlations were statistically significant (all ps<0.001) and reflect moderate agreement (Koo & Li, 2016).

At the item level, Gwet’s AC2 indicated moderate-to-high agreement across age-by-sex subgroups. Weighted κ values were generally in the moderate range, although some individual items showed lower estimates. Given the concentrated and skewed response distributions observed for several items, particularly HI Items 11-14, Gwet’s AC2 was given greater interpretive weight because it is less affected by marginal response distributions (see discussion of kappa’s paradox above). Overall, the subgroup results indicated moderate-to-high parent-youth agreement across age and sex groups.

The relationships between severity of depression and ADHD and indices of agreement could clarify variability in agreement across youth. Given the limitations inherent in using regression to examine relationships with difference scores to address this issue (Bland & Altman, 1995; Laird & De Los Reyes, 2013), we conducted traditional Bland-Altman analyses to examine parent-adolescent agreement in ADHD symptom ratings and modified Bland-Altman analyses to examine whether agreement varied as a function of depression severity. Traditional Bland-Altman plots display the difference between parent and adolescent ratings as a function of the average of the parent and adolescent ratings, (see Figure 1) whereas modified Bland-Altman plots display the difference between parent and adolescent ADHD ratings as a function of depression severity (see Figure 2). Thus, a difference score of zero indicates exact agreement between informants, negative values indicate that parents reported fewer symptoms than adolescents, and positive values indicate that parents reported more symptoms than adolescents. Results indicated minimal average disagreement between parent and adolescent ratings for both symptom domains. For IA, the mean difference was −0.10 points, indicating that parents rated inattention symptoms on average, 0.10 points lower than adolescents, with 95% limits of agreement −9.69 to 9.4. For HI, the mean difference was −0.44 points, indicating that parents rated HI symptoms, on average 0.44 points lower than adolescents with 95% limits of agreement ranging from −7.90 to 7.03. Thus, although average differences were minimal for both symptom domains, the relatively wide limits of agreement indicate substantially greater individual differences between informants than suggested by the mean differences alone.

**Figure 1.**
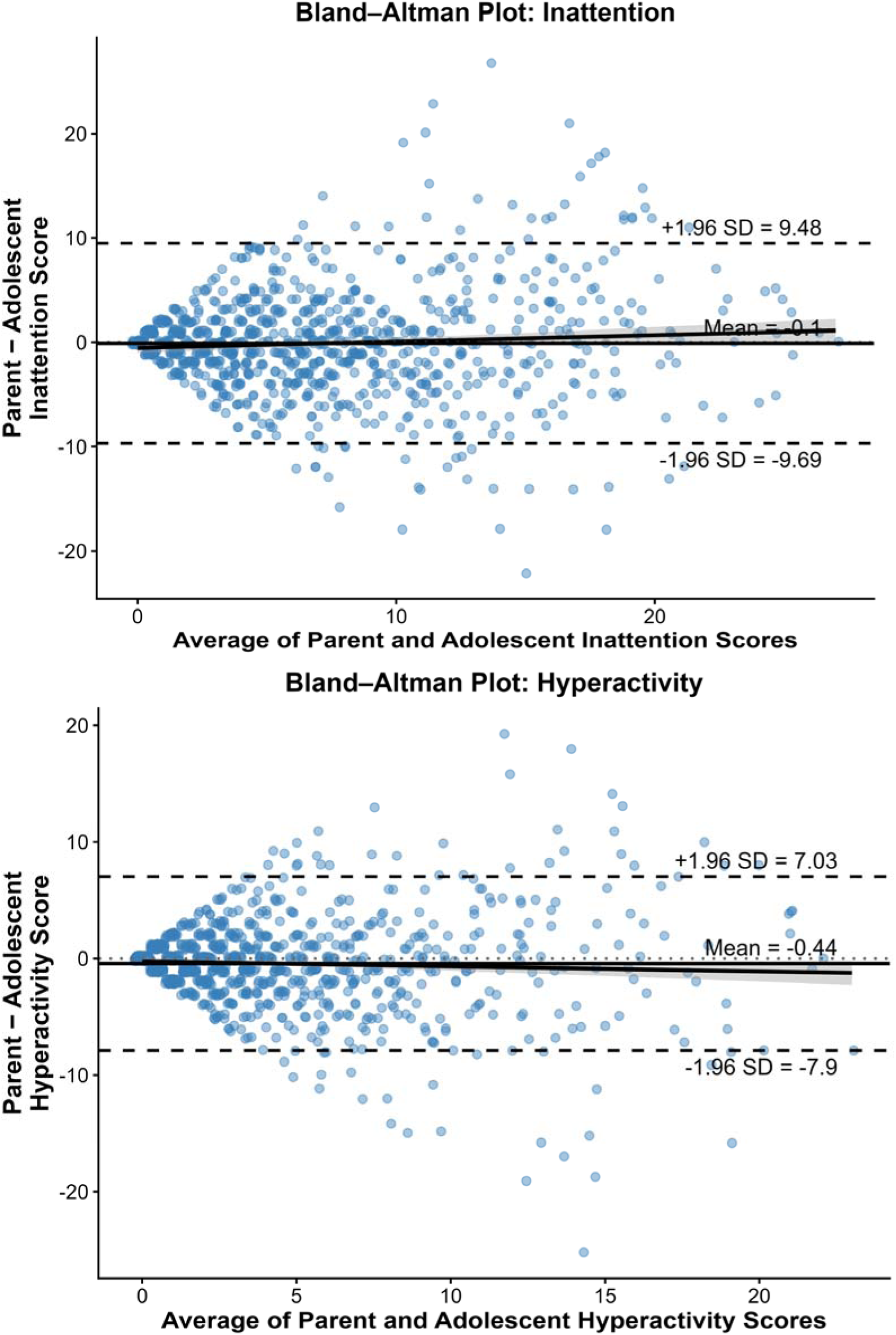
Results of Bland Altman analyses of parent-rated IA and HI symptoms. *Note.* Bland-Altman plots show the average parent and adolescent ratings on the x-axis and the difference between ratings (parent-adolescent) on the y-axis. The solid line indicates the mean difference, and dashed lines indicate the 95% limits of agreement. Negative values indicate lower parent ratings relative to adolescent ratings.

**Figure 2.**
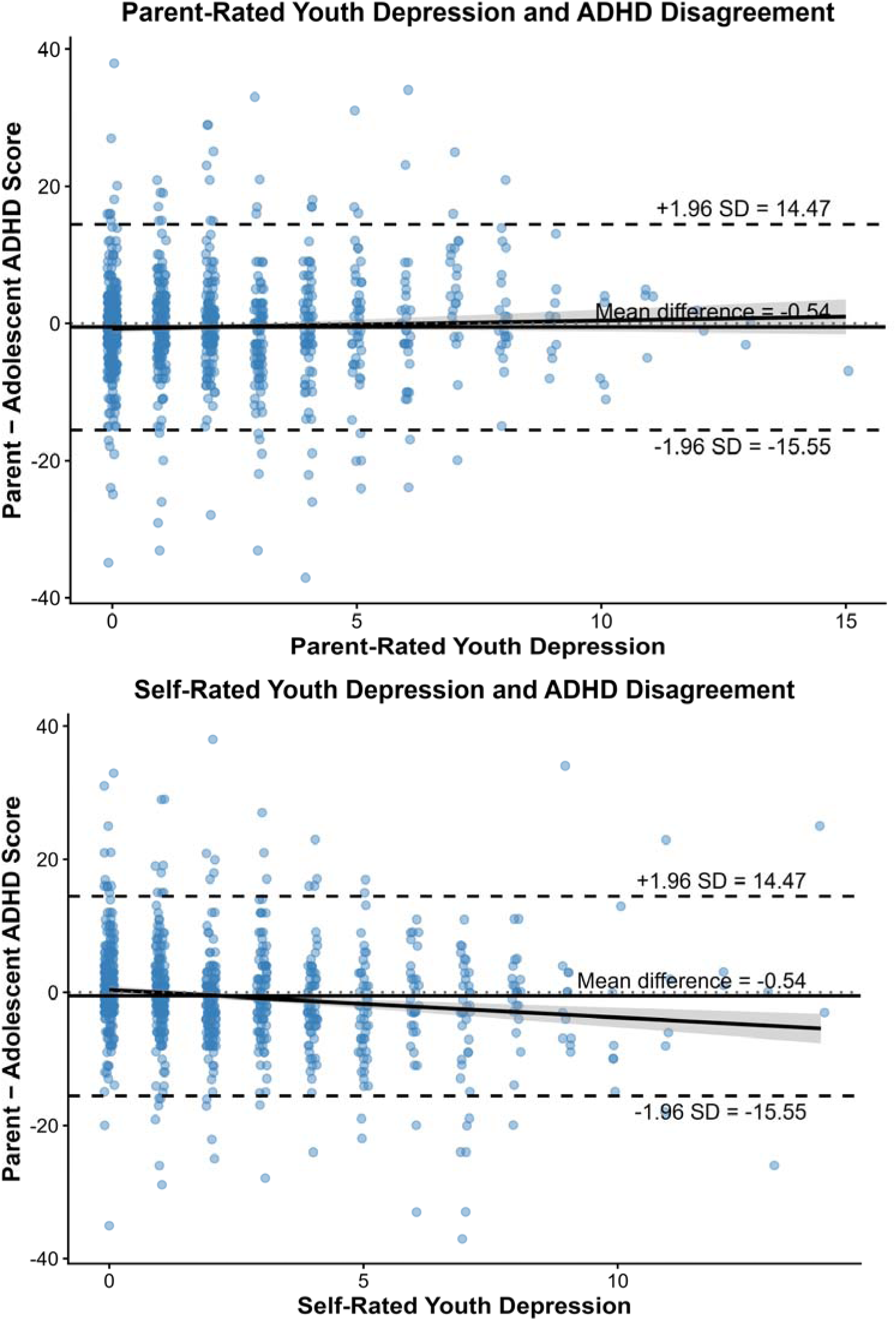
Results of Modified Bland Altman analyses of parent and self-rated depression symptom. *Note.* Modified Bland-Altman plots display parent-adolescent discrepancies in ADHD symptom ratings as a function of depression severity. The solid line indicates the mean difference, and dashed lines indicate the 95% limits of agreement. Negative values indicate lower parent ratings relative to adolescent ratings.

Visual inspection of the traditional Bland-Altman plots suggested increasing variability in parent-adolescent discrepancies as average ADHD symptom severity increased, particularly at higher levels of IA and HI symptoms. This pattern is consistent with potential heteroscedasticity and poorer agreement among participants with greater ADHD symptom severity.

Visual inspection of the modified Bland-Altman plots indicated little evidence of systemic bias as a function of parent-rated depression severity. However, a modest negative trend was observed when discrepancies were plotted against adolescent self-rated depression severity, suggesting that as adolescents reported greater depressive symptoms, parents tended to report relatively fewer ADHD symptoms compared to their adolescents. In addition, variability in discrepancy scores appeared somewhat greater at higher levels of depression severity, particularly for self-reported depression symptoms, suggesting poorer agreement among youth with more severe self-reported depressive symptoms.

## Discussion

### Agreement and Associated Characteristics

Overall absolute agreement between factor scores and total scores on the VAS ratings of ADHD symptoms by parents and adolescents was in the moderate range (ICCs = 0.67 to 0.71) and rank order agreement was moderate to strong. Item level agreement was high across the entire sample (range 0.79-0.95; *M*=0.85; *SD*=0.05) with little variability between items. Q-correlations revealed that parents and adolescents demonstrated low agreement on relative strengths and weaknesses within a factor. This was especially true for IA symptoms. Nevertheless, these results suggest that using most traditional metrics of agreement the VAS has good inter-rater reliability with adolescents and their parents. However, there is evidence in the literature that adolescents with ADHD tend to under-report their symptoms including one study in which almost a quarter of adolescents with ADHD denied any symptoms (Sibley et al., 2019). Additionally, there is evidence that other factors such as age, sex, and levels of depression reduce agreement. Thus, despite good agreement across the population of adolescents, there are factors that appear to diminish agreement.

#### Age and Sex

Results of this study are inconsistent with age and sex impacting levels of agreement. There were negligible differences in agreement statistics (Gwets AC2) between multiple age and sex bands within the sample (IA items Gwets AC2 range 0.74-0.88; *M*=0.82; *SD*=0.03; HI items range 0.77-0.97; *M*=0.89; *SD*=0.05). Adolescents in this sample ranged from 12 years of age to 18 years of age and some previous studies showing age effects included younger children (age range in Hemmingsson et al., 2017 systematic review ranged from 5 to 19 years of age). As a result, age differences in agreement may not have been apparent in our study due to not including children younger than 12 years old. Previous studies showing a sex difference on levels of parent adolescent agreement used data from adolescents diagnosed with ADHD (e.g., Frick et al., 2025), in contrast to the community sample used in the present study. Thus, sex differences in cross-informant discrepancies may exist for youth diagnosed with ADHD, but not across the broader population of adolescents.

#### Depression Symptoms

In this study using a large community sample, self-ratings of depression were related to parent-adolescent agreement (see Figure 2). When adolescents rated depression, higher levels of depression were associated with lower levels of agreement. Specifically, adolescents’ ratings were increasingly higher than parent ratings of ADHD symptoms as the severity of depression increased, consistent with the negative self-appraisals common among youth with depression. This trend is first apparent at fairly low levels of self-reported depression on the VQIDS. Jiang and Johnston (2014) hypothesized that the overestimation of competencies common in youth with ADHD may be offset among youth with comorbid depression yielding greater agreement with parents on ratings of ADHD symptoms than youth with only ADHD. Our results cannot evaluate that hypothesis; however, the evidence of negative self-appraisals applying to ratings of ADHD symptoms is consistent with that hypothesis.

#### ADHD Symptoms

Much of the literature on parent-adolescent agreement on ratings of ADHD symptoms is conducted with samples of youth diagnosed with ADHD. Although some report substantial disagreement (e.g., almost a quarter of adolescents diagnosed with ADHD denied any ADHD symptoms, Sibley et al., 2019), Sibley and colleagues (2019) also reported that approximately one-third of parent-adolescent dyads agreed on the adolescent’s level of ADHD symptoms. The results of our study are consistent with the finding of greater disagreement when teens have higher levels of ADHD compared to lower levels (see Figure 1). The Bland Altman analyses examining agreement in relation to average level of parent and adolescent ratings of IN and HI show that agreement diminishes when scores on these factors are between 10 and 15 (approximately greater than the 75th percentile; Evans et al., 2026). Although the levels of disagreement are not large, the fact that disagreement increases at the low end of the clinical range is consistent with the findings from studies with samples restricted to adolescents with ADHD.

### Kappa’s Paradox

The results of our study provide another example of kappa’s paradox. As expected, many of the HI symptoms had the lowest rates of endorsements for adolescents. Thus, these items have the most skewed distributions of the 18 items on the VAS. They are also the items that among the oldest groups of adolescents have the lowest κ (see bottom rows of Table 5). In contrast, Gwet’s AC2 indices remain strong for those age groups and items and follow a pattern similar to percent agreement. This suggests that studies using κ as the measure of agreement are likely to underestimate the level of agreement for items with considerable skewness and low rates of endorsement. It is common for some items on a variety of measures of psychopathology to include items that will have very low endorsement, especially for some populations. As a result, based on our results we encourage those conducting studies of inter-rater agreement at the item-level with scales assessing psychopathology to use the Gwet AC2 index of agreement instead of κ.

### Limitations

The results of this study address the primary research questions pertaining to inter-rater reliability and the VAS. Much of the research on the inter-rater agreement of ADHD symptoms was completed with samples of youth diagnosed with ADHD. Although this research informs the present study, the results of this study do not add to that literature. In addition, although we used data from a diverse and large sample, it is not completely representative of the population in the United States. Some minorities are under-represented and college-educated parents are over-represented.

### Clinical Implications

The results of this study provide evidence for the growing literature supporting the psychometric properties of the VAS (Anderson et al., 2022; Evans et al., 2026). Severity of ADHD symptoms and depression symptoms emerged as characteristics of adolescents associated with levels of disagreement. Within the age range of participants in this study (i.e., 12 – 18) there was no relationship between age and sex on levels of agreement. Based on these findings and other literature, clinicians may expect disagreement when ADHD symptoms and/or depression symptoms are high. Although multiple indices of agreement support moderate or greater inter-rater reliability, there is poor agreement pertaining to the relative rankings of ADHD symptoms within each factor. Thus, clinicians should not assume agreement between parents and adolescents on the profiles of symptoms within each factor (relative strengths and weaknesses). As with any discrepancies in reporting between parents and adolescents, clinicians are encouraged to explore the unique perspectives of each rater to the extent necessary to inform diagnosis or treatment.

Although these findings are encouraging and give users of the VAS confidence in administering and interpreting the scale, there is clearly room for improvement. Johnston and colleagues (2014) reported evidence that providing education to parents about the meaning of items on ADHD symptom rating scales (like the VAS) improves accuracy. Although current wording of the items makes the scale easy and quick to read and complete, revisions to the wording that enhance clarity may improve the validity and reliability of the measure.

## Data Availability

All data produced in the present study are available upon reasonable request to the authors.

## Acknowledgements

Supported by gifts from the Chlapaty, DiMarco, and Zipfel families to Nationwide Children’s Hospital to support the Institute for Mental and Behavioral Health Research (IMBHR).

## Disclosures

Dr. Evans is the Editor-in-Chief of the journal *School Mental Health* and receives royalties from Springer, Oxford Press, and Taylor & Francis publishing. Dr. Youngstrom is the co-founder and Executive Director of Helping Give Away Psychological Science, a 501c3; he received royalties from the American Psychological Association and Guilford Press, and he holds equity in Joe Startup Technologies.

The data that support the findings of this study are available from the corresponding author, SWE, upon reasonable request.

